# Stress-related emotional and behavioural impact following the first COVID-19 outbreak peak

**DOI:** 10.1101/2020.12.08.20245787

**Authors:** Asaf Benjamin, Yael Kuperman, Noa Eren, Maya Amitai, Hagai Rossman, Smadar Shilo, Tomer Meir, Ayya Keshet, Orit Nuttman Shwartz, Eran Segal, Alon Chen

## Abstract

**Background:** The coronavirus disease 2019 (COVID-19) pandemic poses multiple psychologically-stressful challenges and is associated with increased risk for mental illness. Previous studies have mostly focused on the psychopathological symptoms associated with the outbreak peak.

**Methods:** We examined the behavioural and mental health impact of the pandemic in Israel using an online survey. We collected 12,125 responses from 4,933 adult respondents during six weeks encompassing the end of the first outbreak and the beginning of the second. We used clinically validated instruments (Brief Symptom Inventory 18 (BSI-18), Perceived stress scale (PSS), Brief COPE inventory) to assess anxiety- and depression-related emotional distress, symptoms, and coping strategies, as well as questions designed to specifically assess COVID-19-related concerns.

**Results:** Respondents indicated worrying more about the situation in their country and their close ones contracting the virus, than about their own health and financial situation. The reported distress correlates with the number of new COVID-19 cases and higher emotional burden was associated with being female, younger, unemployed, living in low socioeconomic status localities, encountering more people, and experiencing physiological symptoms. Unexpectedly, older age and having a prior medical condition were associated with reduced emotional distress.

**Conclusions:** Our findings show that inequalities in mental-health burden associated with the COVID-19 pandemic are relevant also following the initial outbreak, and highlight the environmental context and its importance in understanding individual ability to cope with the long-term stressful challenges of the pandemic.

## Introduction

As of October 31^st^ 2020, the severe acute respiratory syndrome coronavirus 2 (SARS-CoV-2), which causes the coronavirus disease 2019 (COVID-19), has infected 45 million people and taken the lives of over 1,180,000 in more than 200 countries and territories around the world^1^. The pandemic has dramatically affected virtually all aspects of our lives: It has led to the largest global recession since the Great Depression^2^, and to extreme social isolation due to changes in educational and work activities, local lockdowns, and international travel restrictions. Social isolation and financial instability, together with fear of contracting COVID-19 and uncertainty of the future, pose substantial psychological stressors for the general population^3^. It is likely that the pandemic induces a considerable degree of fear, worry and concern in the general population.

Studies of the impact of this pandemic on people’s mental health, and into ways of mitigating adverse mental health consequences for vulnerable subgroups, are critically needed. So far, most of the work in this area has addressed the acute mental health impact of the COVID-19 pandemic, measured during the outbreak peak. Indeed, despite geographical and cultural differences, several studies have provided largely consistent results in these aspects: A study conducted on Chinese residents found that 54% of 1,210 respondents rated the psychological impact of the COVID-19 outbreak as moderate or severe^4^. In an Australian study using an online survey during the outbreak peak, 78% of 5,070 respondents reported worsening of their mental health since the outbreak^5^. An additional study performed on a Spanish cohort (n = 3,840), found that age, economic stability, and being male, were all negatively correlated with reports of depression, anxiety, and PTSD during the initial stage of the COVID-19 pandemic^6^. These and other studies describe the anxiety induced by the pandemic and its negative correlation with sleep quality and social support, both in the general population and in susceptible groups, such as healthcare staff members^7–9^ and self-isolated people^10,11^.

To further shed light on this emerging global picture, we set out to assess the long-term mental health-related effects of the pandemic on the adult population in Israel using an online survey. During the six weeks between April 28^th^ and June 9^th^ 2020, we collected 12,125 responses from 4,933 adult respondents (see study population description in *Methods* section). The respondents agreed to answer a two-stage online questionnaire, where in the first stage, they reported on COVID-19-related physiological symptoms and behaviours together with background demographic and medical information, and in the second stage on the effects of COVID-19 on their psychological and emotional state (see *Methods*). Importantly, our data were collected after the initial outbreak peak, and thus reflect a period in which people have had the chance to adapt to the new circumstances, rather than the initial reactions to the outbreak. These six weeks of data collection allowed us a broad and dynamic view of the period between the first and the second outbreaks.

## Results

### COVID-19-linked stressors induced mainly non-self-centred concerns

While recent studies are starting to address the psychological and emotional effects of the pandemic, less is known regarding the underlying motives. Here, we examine the specific reasons that may underlie the psychological and emotional effects of the pandemic. Thus, to try and assess what people are most worried about during the pandemic, we asked respondents to rate the extent to which they were concerned about specific issues. Despite the many unknowns about the COVID-19 disease and its potential effects on our personal lives in the future, respondents reported lower levels of concern about their personal situation - namely, contracting the coronavirus and their financial situation - than they did about the situation in their country and about people close to them contracting the virus (Fig.1e-f; Friedman’s test with *post-hoc* Bonferroni correction for multiple comparisons: n=4,882; Chi^2=4,971; df=4; for all pairwise comparisons: p<1e-8).

**Figure 1.**
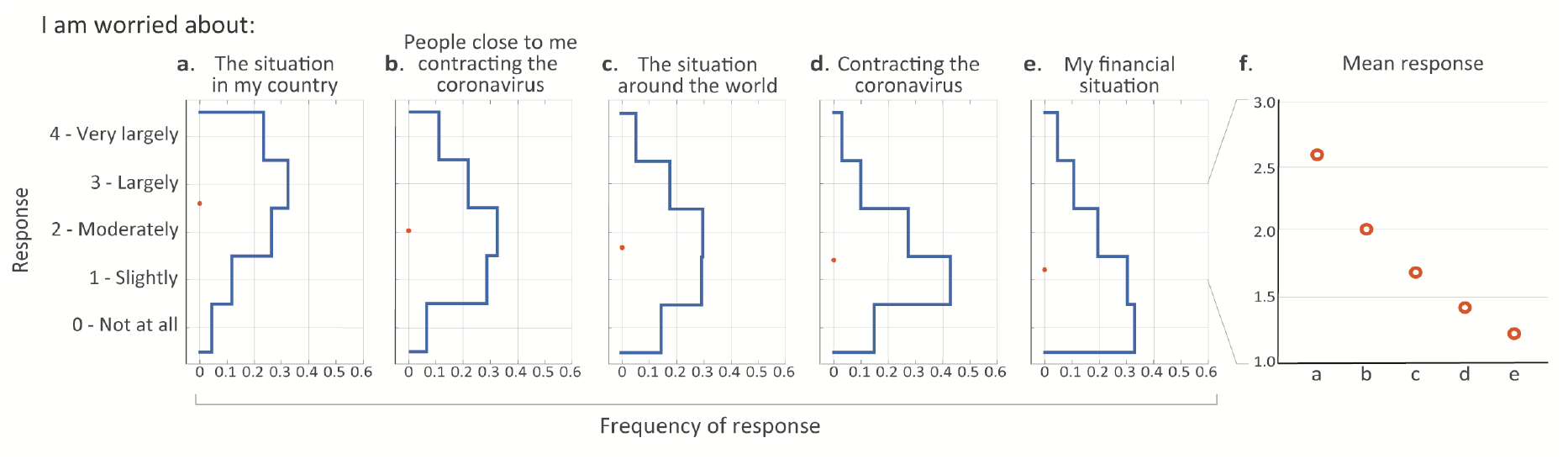
COVID-19 induced mainly non-self-centred concerns. (a-e) Blue lines represent distributions of responses for specific reasons for worry among all respondents. Orange circles represent response means. (f) Zoomed-in view of the response means shown in panels a-e. Note that all five SE ranges are shorter than the circle diameters and were thus omitted from the plot.

In addition to specific reasons for worry, we looked into reports of distress-related symptoms and emotions more generally, using questions from the anxiety and depression subscales of the brief symptom inventory 18 (BSI-18^12^) and from the perceived stress scale (PSS^13^) (Supp. Fig 1). In our cohort, the mean BSI-18 scores for anxiety and depression were 0.72±0.8 and 0.42±0.61, respectively. These scores are similar to the Israeli norm (anxiety: 0.85±0.71; depression: 0.7±0.69), which is based on a nationwide representative sample of 510 community respondents between the ages of 35 and 65 years^14^. We further asked about experiencing several stress-related physiological symptoms and about the stress-coping strategies used (part of the brief-COPE questionnaire^15^). To gain a more integrative and concise description of respondents’ emotional responses, we examined to what extent answers were correlated across questions and could thus be compactly represented by a smaller number of factors. We used factor analysis on the answers to the distress- and worry-related questions, which revealed three principal factors or components (see *Methods* and Supp. Fig. 2). The three factors respectively corresponded to questions related to: (1) general emotional distress and worrying about your financial situation (henceforth: “general emotional distress”); (2) worrying about contracting COVID-19 and about people close to you contracting COVID-19 (henceforth: “COVID-19 infection concern”), and (3) worrying about the situation in Israel and around the world (henceforth: “national and global concern”). In most of the following analyses, we focus on these three measures, along with the number of stress-related physiological symptoms experienced and the number of stress-coping strategies used.

### Reported distress correlates with the number of new COVID-19 cases

Next, we examined whether and how responses changed with time. We observed qualitatively similar characteristics between the temporal dynamics of our five main scores and the number of new daily COVID-19 cases as published by the Israeli Ministry of Health (health.gov.il; Fig. 2a-e). All of these scores seem to gradually decline over the first several weeks, together with the decline in the numbers of new daily COVID-19 patients. Similarly, around May 26^th^, as the numbers of new cases started to rise, so did the reports. However, about a week later, despite the continued rise in new cases, these values started to decline, presumably reflecting an adaptation or behavioural habituation to this new situation. Thus, we quantified the correlation between the number of new cases and these five scores, and found a statistically significant correlation for the general emotional distress scale (Kendall’s correlation coefficient with Bonferroni correction for multiple comparisons: Fig. 2a; n=4,841; Tau=0.03; p=0.009), the number of stress-related symptoms (Fig. 2d; n=4,933; Tau=0.03; p=0.0186) and the number of stress coping strategies used (Fig. 2e; n=4,933; Tau=0.11; p<1e-21), but not for the COVID-19 infection concern (Fig. 2b; n=4,841; Tau=0.01; p=0.733) and the national and global concern (Fig. 2c; n=4,841; Tau=-0.01; p=0.963). Notably, although the dynamics of the national-global concern seem somewhat similar to those of the other scales (a decrease followed by an increase and another decrease), its initial rise starts earlier and it seems largely more stable compared to the other scales (Fig. 2c). Overall, these correlations may demonstrate our questionnaire’s sensitivity in capturing stress-related effects of COVID-19 in real-time. Interestingly, a similar correlation between stress symptoms (assessed by social media data mining) and the number of new COVID-19 cases was also found in a study in the United States^16^.

**Figure 2.**
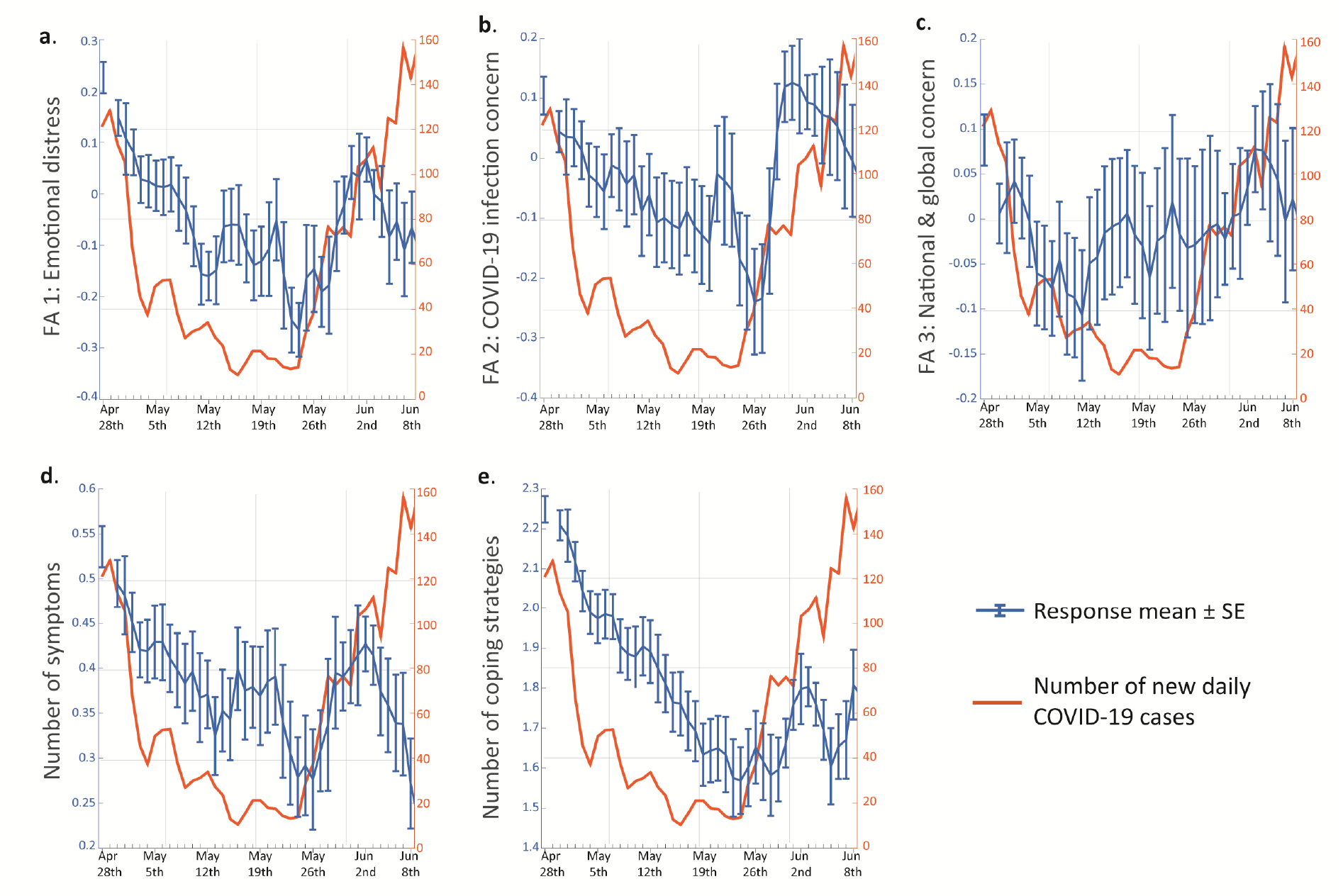
The temporal dynamics of reported distress correlate with the number of new COVID-19 cases. (a-e) Daily means and SE of (a) emotional distress, (b) COVID-19 infection concern, (c) national and global concern, (d) number of stress-related symptoms and (e) number of stress-coping strategies. Orange curves indicate the number of newly diagnosed COVID-19 patients as published by the Israeli Ministry of Health (www.health.gov.il). Kendall’s correlation coefficient with Bonferroni correction for multiple comparisons: (a) Tau=0.03; p=0.009); (b) Tau=0.01; p=0.733; (c) Tau=-0.01; p=0.963; (d) Tau=0.03; p=0.0186); (e) Tau=0.11; p<1e-21).

### Women report higher levels of distress than men

Next, we asked whether the emotional response to the pandemic differed between genders (see note regarding non-binary genders in *Methods* section). Similar to previous studies^17,18^, we found that female respondents reported higher levels of distress on the general emotional distress scale (Fig. 3a-b; Mann-Whitney U test; n_M_=2,178, n_F_=2,473; U=0.55; p=1.7e-9); the COVID-19 infection concern scale (Fig. 3a,c; n_M_=2,178, n_F_=2,473; U=0.55; p<4.5e-9); and the national-global concern scale (Fig. 3a,d; n_M_=2,178, n_F_=2,473; U=0.52; p<0.024). In accordance with the higher levels of emotional distress, we also found that women reported experiencing a greater number of stress-related symptoms (Fig. 3a,e; n_M_=2,212, n_F_=2,525; U=0.56; p=2e-14), and using more stress-coping strategies (Fig. 3a,f; n_M_=2,212, n_F_=2,525; U=0.56; p=4e-13). Specifically, women were more likely to report experiencing increased heart rate (Supp. Fig. 3; n_M_=2,212, n_F_=2,525; see per-question contingency table counts in figure; Fisher’s exact test: odds ratio (OR)=1.69; p=0.0026), increased appetite (Supp. Fig. 3; OR=1.56; p=0.0006) and trouble sleeping (Supp. Fig. 3; OR=1.44; p=7.2e7). Women were also more likely to report using specific coping strategies, such as contacting someone for support (Supp. Fig. 3; OR=2.79; p<1e-26) and exercising or meditating (Supp. Fig. 3; OR=1.19; p=0.0434). This may suggest that more coping methods are needed to alleviate a greater sense of emotional distress. Importantly, considering the full distribution of responses, the differences between genders seem much more prominent at the lower levels of general emotional distress, as well as for lower numbers of symptoms and coping strategies (Fig. 3a,d-e), while they seem less prominent for both high and low levels of concern about COVID-19 and about the national and global situation (Fig. 3b-c).

**Figure 3.**
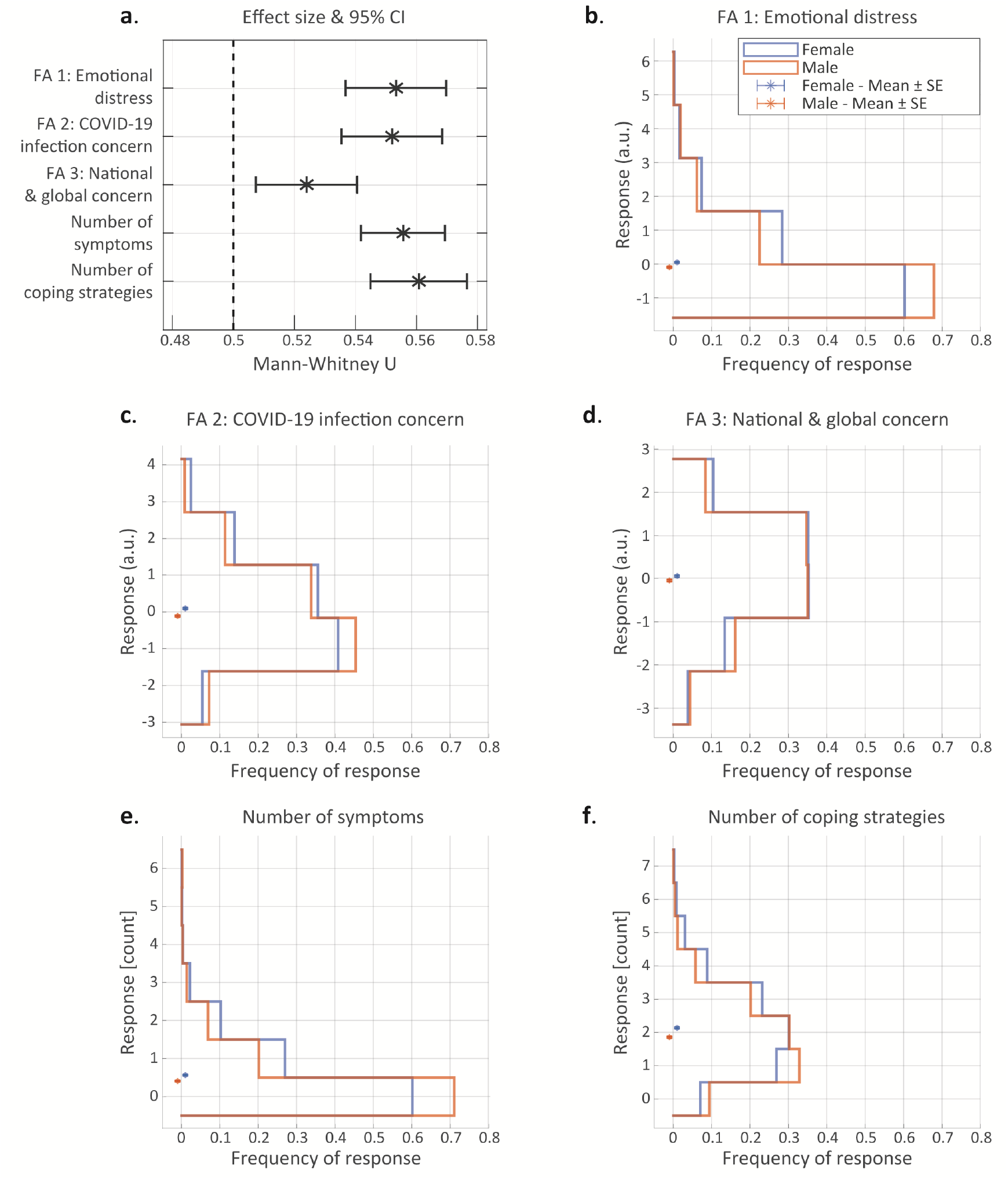
Women report higher levels of distress than men. (a) Effect sizes and 95% confidence intervals of the Mann-Whitney U statistic for female vs. male responses on the five stress-related scales. (b-f): stress-related response distributions of women (blue) and men (orange): (b) emotional distress scale; (c) COVID-19 infection concern scale; (d) national-global concern scale; (e) number of stress-related symptoms; (f) number of stress-coping strategies. Asterisks in b-f represent scale means.

### Age negatively correlates with reported distress

Since age is a known factor influencing the stress response^19^, we evaluated the effect of age on people’s emotional responses to the pandemic. We quantified the correlation between respondents’ age and their answers, and found that younger respondents scored significantly higher on the general emotional distress scale (Fig. 4a-b; Kendall’s correlation coefficient with Bonferroni correction for multiple comparisons: n=4,641; Tau=-0.2; p<1e-86), and on the COVID-19 infection concern scale (Fig. 4a,c; n=4,641; Tau=-0.11; p<1e-25). This reduction in concern with age may seem counterintuitive, in light of the significantly increased risk for complications among older COVID-19 patients^20^, but it is consistent with several previous studies^6,11,21^.

**Figure 4.**
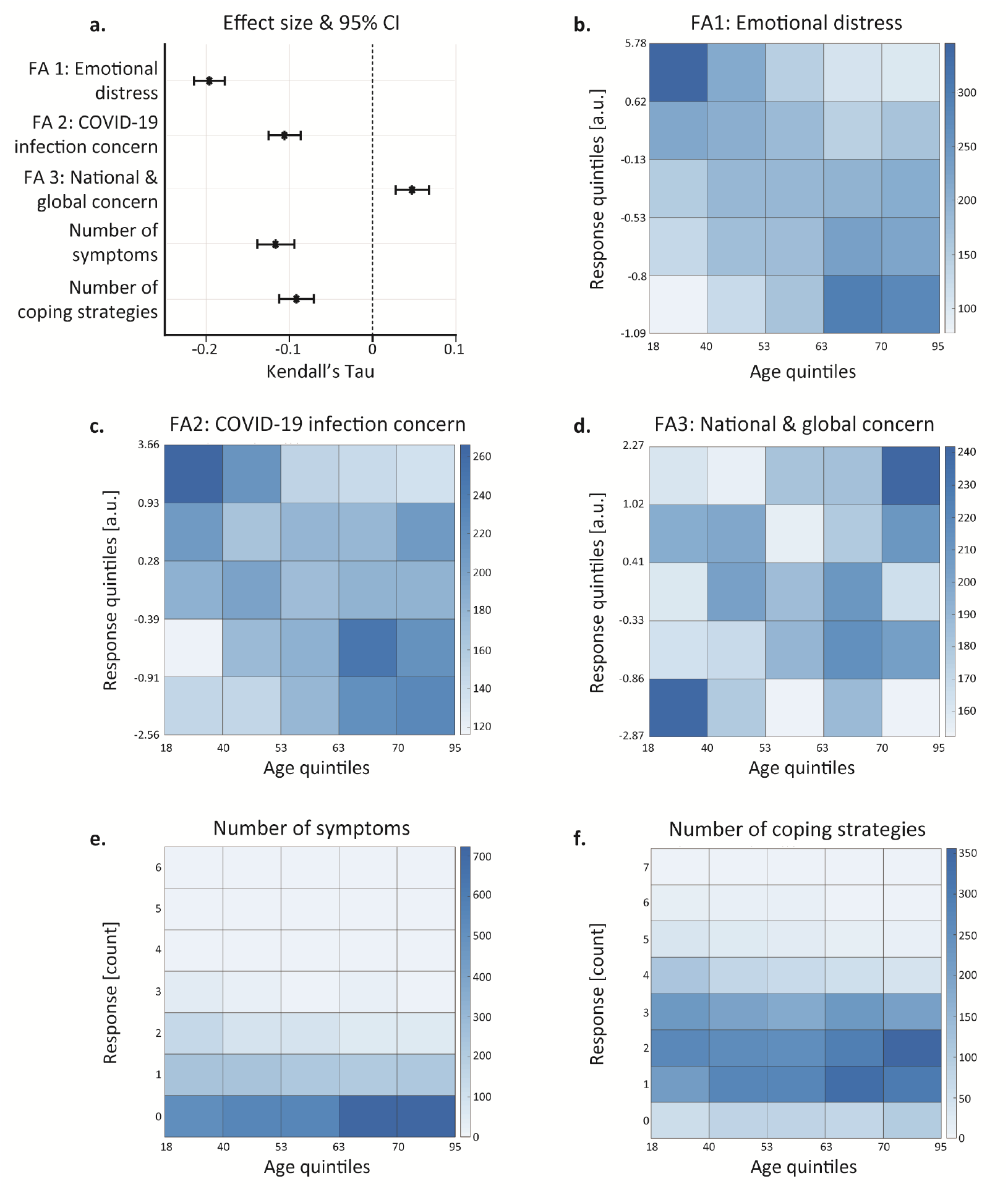
Age negatively correlates with reported distress. (a) Effect sizes and 95% confidence intervals of Kendall’s Tau for the association between age and responses on the five stress-related scales. (b-f) Heatmaps representing frequency of responses for each age-response subgroup. Age and the three continuous response scores (b-d) were divided into five equally sized subgroups based on quintiles, while the (integer) number of symptoms and coping strategies were left as is (e-f; see methods). Kendall’s correlation coefficient with Bonferroni correction for multiple comparisons: (b) n=4,641; Tau=-0.2; p<1e-86; (c) n=4,641; Tau=-0.11; p<1e-25; (d) n=4,641; Tau=0.05; p=8.38e-06; (e) n=4,728; Tau=-0.12; p<1e-22; (f) n=4,728; Tau=-0.09; p<1e-15.

Age was also negatively correlated with the number of stress-related symptoms reported (Fig. 4a,e; n=4,728; Tau=-0.12; p<1e-22) and with the number of stress-coping strategies used (Fig. 4a,f; n=4,728; Tau=-0.09; p<1e-15). Importantly, despite a lower total number of coping strategies, older respondents exhibited an increased tendency to exercise and/or meditate (Supp. Fig. 4; Mann-Whitney U test: n=4,728; U=0.58; p<1e-21). Intriguingly, in contrast with all other types of concern and distress, older respondents scored higher on the national-global concern scale (Fig. 4a,d; n=4,641; Tau=0.05; p=8.38e-06), supporting the separation of these concerns into discrete factors.

To visualize these associations, we divided the respondents into five equally sized subgroups according to their age, and divided the main continuous response scores (for the general emotional distress, COVID-19 infection concern, and national-global concern scales) into five subgroups according to the responses. We then plotted the frequency of each pair of age-response groups (Fig. 4b-f; see *Methods* for details). Under the null hypothesis of no association between respondents’ age and their responses, each matrix cell is expected to have the same frequency, a stark contrast to the matrices we obtained (Fig. 4b-f). Taken together, these analyses reveal that the pandemic may be affecting younger people’s mental and emotional states more severely.

### Employment status is associated with reported distress

As the pandemic influenced the stability of working places^22–24^, we assessed the association between respondents’ employment status and their emotional response to the pandemic. We divided the respondents into four groups: (1) respondents who are currently working (n=2,546); (2) respondents who lost their job due to COVID-19 (either termination of employment (ToE), or on paid or unpaid leave, or forced retirement; n=700); (3) respondents who were unemployed since before the pandemic (n=405); and (4) respondents who were retired since before the pandemic (n=1,131). Comparing the responses between these four subgroups revealed several interesting findings: First, work status was significantly associated with general emotional distress, COVID-19 infection concern, stress-related symptoms, and coping strategies. National-global concern was the only scale that did not significantly associate with work status (Fig. 5; Kruskal–Wallis test by ranks with post-hoc Bonferroni correction for multiple comparisons; general emotional distress: n=4,697; df=3; Chi^2=236.73; p<1e-10; COVID-19 infection concern: n=4,697; df=3; Chi^2=30.79; p=9.39e-7; national-global concern: n=4,697; df=3; Chi^2=6.92; p=0.074; number of stress-related symptoms: n=4,725; df=3; Chi^2=69.4; p<1e-10; number of stress-coping strategies: n=4,725; df=3; Chi^2=33.6; p-2.4e-7).

**Figure 5.**
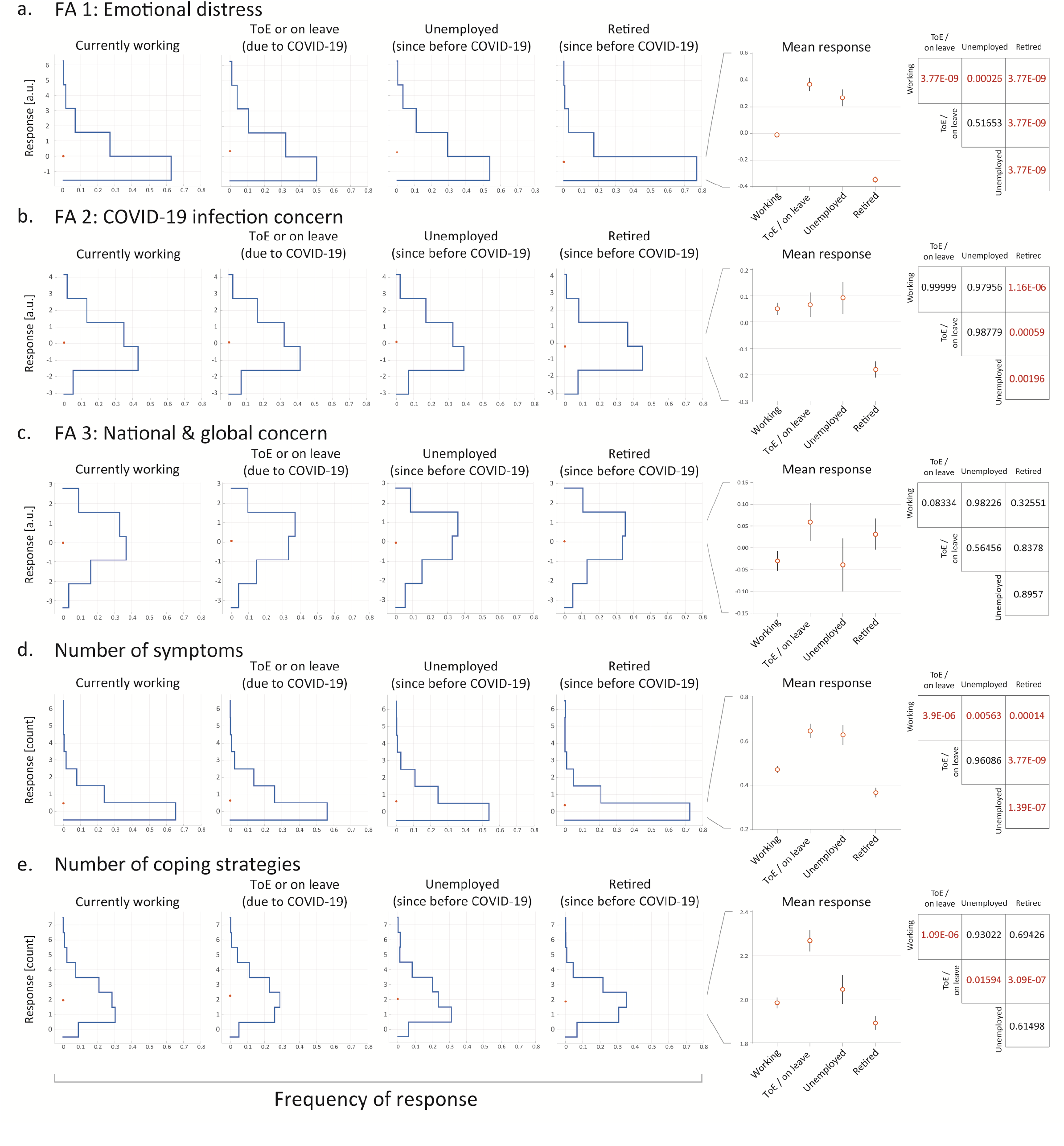
Employment status is associated with reported distress. (a-e) Left: response distributions and means for each of the four work-status subgroups. Centre: Zoomed-in views of the four mean responses are shown for clearer comparison. Right: Post-hoc pairwise comparison p-values with Tukey-Kramer’s honestly significant difference correction for multiple comparisons.

Currently unemployed respondents (both since before and due to the pandemic) reported significantly higher emotional distress and a significantly higher number of stress-related symptoms compared to currently employed respondents (see statistics in Fig. 5a,d). Moreover, respondents who were terminated or on leave due to COVID-19 reported using significantly more stress-coping strategies compared to respondents who were either currently working or unemployed since before the pandemic (see statistics in Fig. 5e). In accordance with our age-related findings, the retired group reported significantly lower levels of emotional distress and concern about COVID-19 infection, and a significantly lower number of stress-related symptoms compared to all the other subgroups (see pairwise comparisons in Fig 5a-b, d). Retired respondents also reported using the lowest number of stress-coping strategies, but this difference was only statistically significant compared with respondents who were terminated or on-leave due to COVID-19 (see statistics in Fig. 5e). Taken together, these results may exemplify the roles employment plays in providing both financial security, social support, and better self-esteem^25^, which ultimately influence emotional state.

### High city socioeconomic status is associated with lower reported distress

Socioeconomic status is a strong predictor of health outcomes, and is generally associated with distress and with prevalence of mental health problems^26^. To evaluate the association between the respondents’ socioeconomic status and their emotional response to the pandemic, and since we did not have details about the respondents’ income, we used their city socioeconomic score (CSS). The CSS is provided by the Israeli Central Bureau of Statistics (cbs.gov.il), who scores all cities, towns and other incorporated settlements in Israel from 1 to 255, with higher numbers corresponding to higher status. We first quantified the correlation between respondents’ CSS and their responses. We found that respondents with lower CSS reported significantly higher levels of emotional distress (Fig. 5a-b; Kendall’s correlation coefficient with Bonferroni correction for multiple comparisons: n=4,009; Tau=-0.041; p=7.9e-4), and using more stress-coping strategies (Fig. 5a,f; n=4,081; Tau=-0.033; p=0.026). More specifically, respondents with lower CSS reported more frequently that they drew strength from belief in god (Supp. Fig. 5; Mann-Whitney U test with Bonferroni correction for multiple comparisons; n=4,088; U=0.29; p<1e-10), and approached a family member or a friend for support (Supp. Fig. 5; n=4,088; U=0.45; p=0.005), but less frequently that they exercised or meditated (Supp. Fig. 5; n=4,088; U=0.54; p=0.001). Moreover, although respondents with lower CSS did not report experiencing more stress-related symptoms overall, they were significantly more likely to report experiencing difficulty breathing (Mann-Whitney U test with Bonferroni correction for multiple comparisons; Supp. Fig. 5; n=4,088; U=0.42; p=0.037). In contrast, higher CSS was positively correlated with the national-global concern scale (Fig. 5a,d; n=4,009; Tau=0.032; p=0.0164) and using exercising or meditating as a coping strategy (Mann-Whitney U test with Bonferroni correction for multiple comparisons; Supp. Fig. 5; n=4,088; U=0.535; p=0.001).

To visualize this relationship in detail, we divided the respondents into five equally sized subgroups according to their CSS, and the continuous responses (general emotional distress, COVID-19 infection concern and national-global concern scales) into five groups according to extent quintiles, and plotted the frequency of each pair of CSS-response groups (fig. 5b-f; see *Methods* for details). Similar to the age plots, under the null hypothesis, we would expect all matrix cells to have a similar frequency, which was not the case here.

### Respondents with prior medical conditions report elevated concern about contracting COVID-19 but reduced general distress

Various medical conditions may increase the risk for complications of COVID-19^27,28^. Therefore, we explored whether having such prior medical conditions influenced respondents’ emotional distress. As expected, respondents with at least one prior medical condition worry more about contracting COVID-19 (Fig. 7a; Mann-Whitney U test with Bonferroni correction for multiple comparisons; n=4,162 respondents reported whether or not they had each of the conditions; n=1,568 respondents had at least one condition; U=0.57; p<1e-10). However, to our surprise, these respondents reported lower levels of distress on the general emotional distress scale (Fig. 7a; U=0.46; p=2.3e-5), were less worried about people close to them contracting COVID-19 (Fig. 7a; U=0.47; p=0.009) and used fewer stress-coping strategies (Fig. 7a; U=0.47; p=8.1e-4).

**Figure 6.**
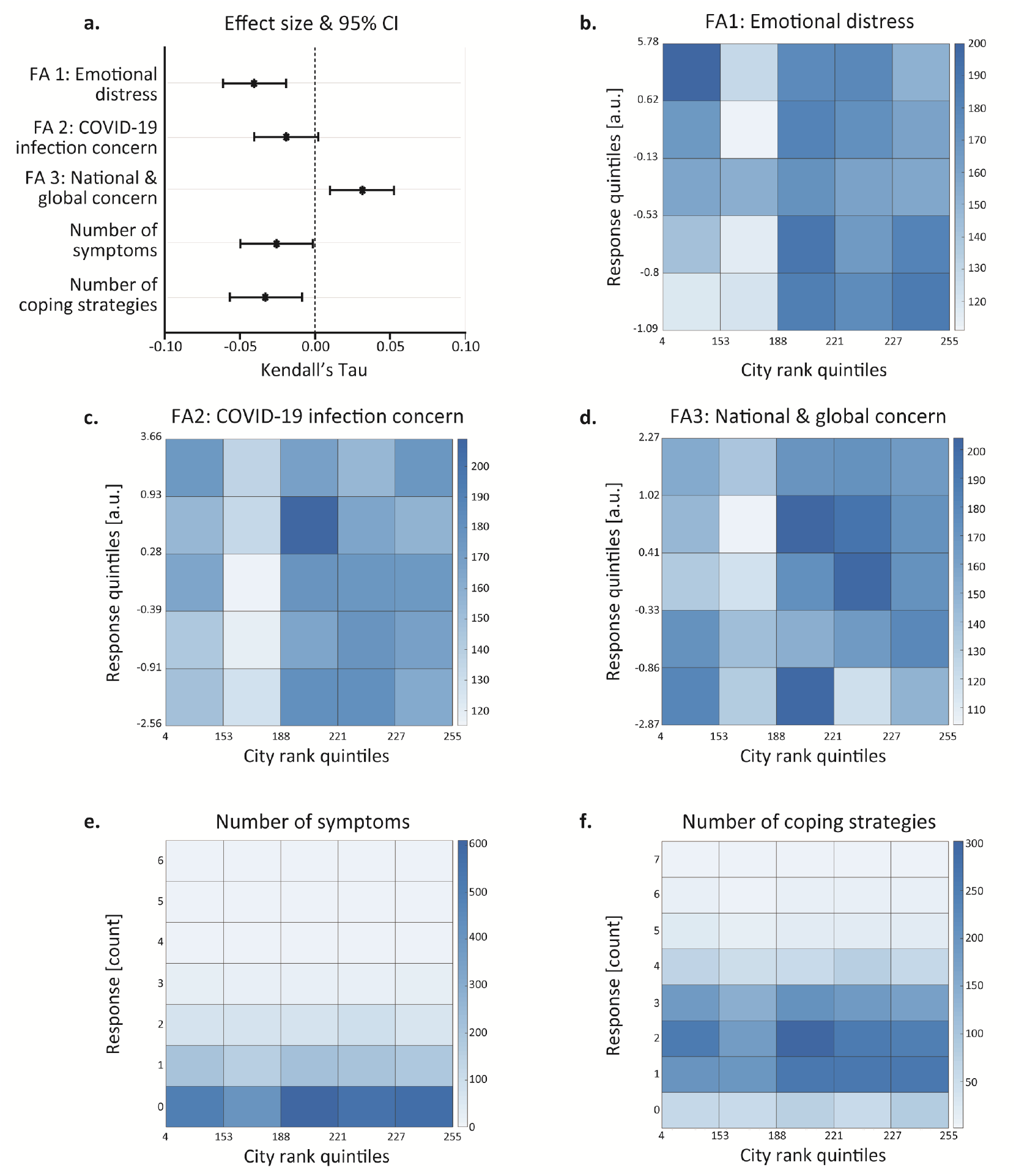
City socioeconomic score (CSS) is associated with reported distress. (a) Effect sizes and 95% confidence intervals of Kendall’s Tau for the association between CSS and responses on the five stress-related scales. (b-f) Heatmaps representing frequency of responses for each CSS-response subgroup pair. CSS and the three continuous response scores (b-d) were divided into five equally sized subgroups based on quintiles, while the (integer) number of symptoms and stress-coping strategies (e-f) were left as is (see methods). Kendall’s correlation coefficient with Bonferroni correction for multiple comparisons: (b) n=4,009; Tau=-0.041; p=7.9e-4; (c) n=4,009; Tau=0.02; p=0.33; (d) n=4,009; Tau=0.032; p=0.0164; (e) n=4,081; Tau=0.026; p=0.191; (f) n=4,081; Tau=-0.033; p=0.026.

**Figure 7.**
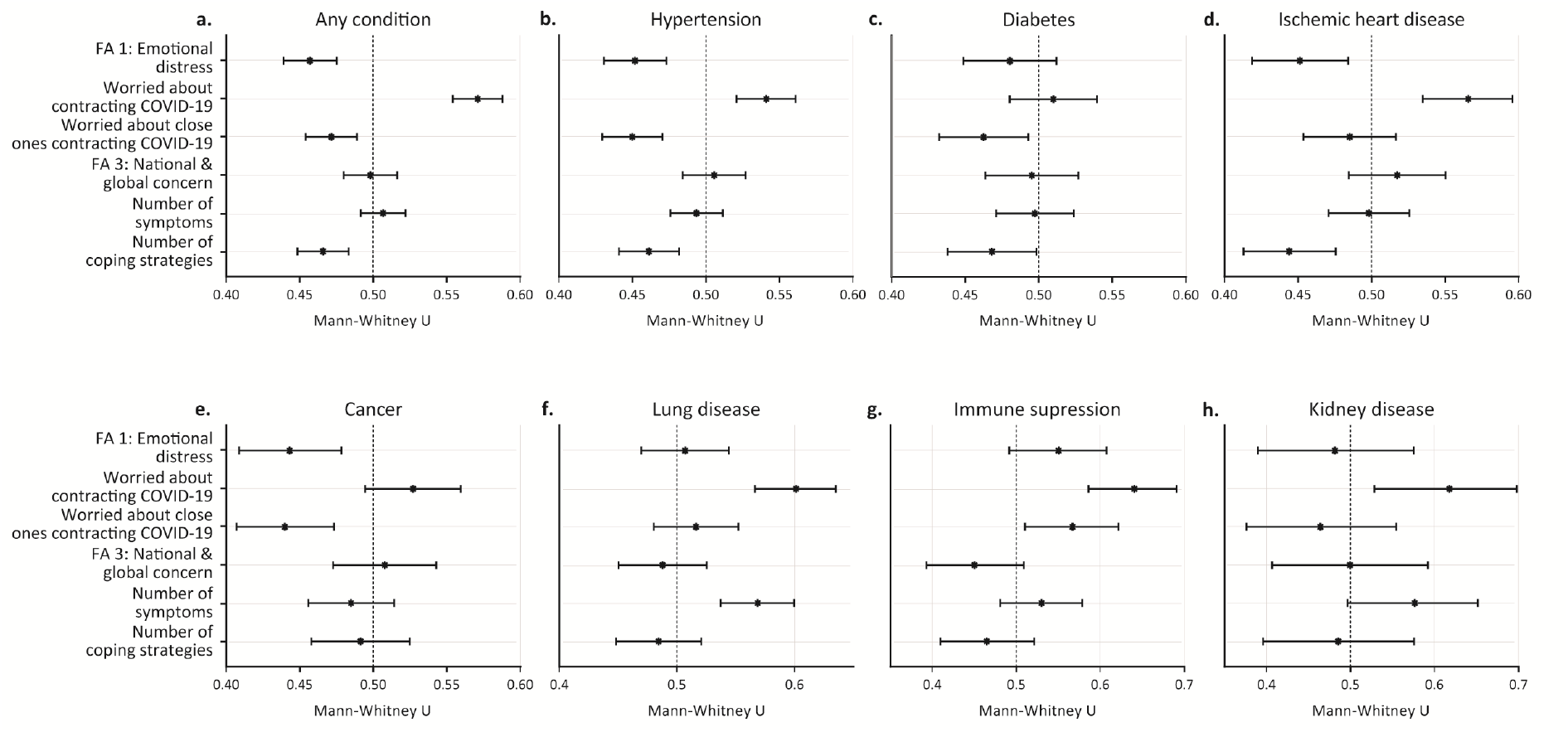
Respondents with prior medical conditions report reduced general distress but elevated concern about contracting COVID-19. (a-h) Effect sizes and 95% confidence intervals of Mann-Whitney’s U for the association between prior medical conditions and respondents’ stress-related responses. Mann-Whitney U test with Bonferroni correction for multiple comparisons.

Next, we examined whether similar findings were consistent across different medical conditions. Indeed, we found a significant elevation in concern about contracting COVID-19 in respondents with hypertension (Fig. 7b; n=907; U=0.54; p=3.7e-4), ischemic heart disease (Fig. 7d; n=324; U=0.56; p=1.8e-4), lung disease (Fig. 7f; n=242; U=0.6; p=1.2e-7), and suppressed immune system (Fig. 7g; n=96; U=0.64; p=4.1e-6), and a nearly-significant elevation in respondents with kidney disease (Fig. 7h; n=36; U=0.62; p=0.058). In contrast, we found lower levels of general emotional distress in respondents with hypertension (Fig. 7b; n=907; U=0.54; p=6e-5), ischemic heart disease (Fig. 7d; n=324; U=0.45; p=0.022) and cancer (Fig. 7e; n=286; U=0.44; p=0.009). Of note, we found no condition associated with reduced worry about contracting COVID-19 or with elevated general emotional distress. We did not find an association between any of these medical conditions and worrying about the national or global situation. Taken together, these analyses demonstrate that people with certain prior medical conditions that might increase the risk for complications of COVID-19 are more worried about contracting COVID-19, but are nevertheless less emotionally distressed in general.

### COVID-19 symptoms and related behavioural factors associate with reported distress

Next, we examined the extent to which activities and symptoms related specifically to COVID-19 were associated with reported distress. These include behaviours or habits meant to reduce one’s risk of contracting COVID-19, such as wearing a mask and/or gloves, and behaviours which increase the risk of contracting COVID-19 through social contact, such as meeting people face to face and using public transport. First, we found that using public transport was associated with reporting significantly higher levels of general emotional distress (Fig. 8a; Mann-Whitney U test with Bonferroni correction for multiple comparisons: n_public_tranport_=140; n_no_public_tranport_=1,378; U=0.62; p=1e-5). Public transport users also showed a statistical trend towards worrying more about contracting COVID-19 themselves (Fig. 8a; U=0.56; p=0.089) and their close ones contracting COVID-19 (Fig. 8a; U=0.56; p=0.071), as well as experiencing more stress-related symptoms (Fig. 8a; U=0.57; p=7.2e-3) and using more stress-coping strategies (Fig. 8a; U=0.59; p=6.6e-4).

**Figure 8.**
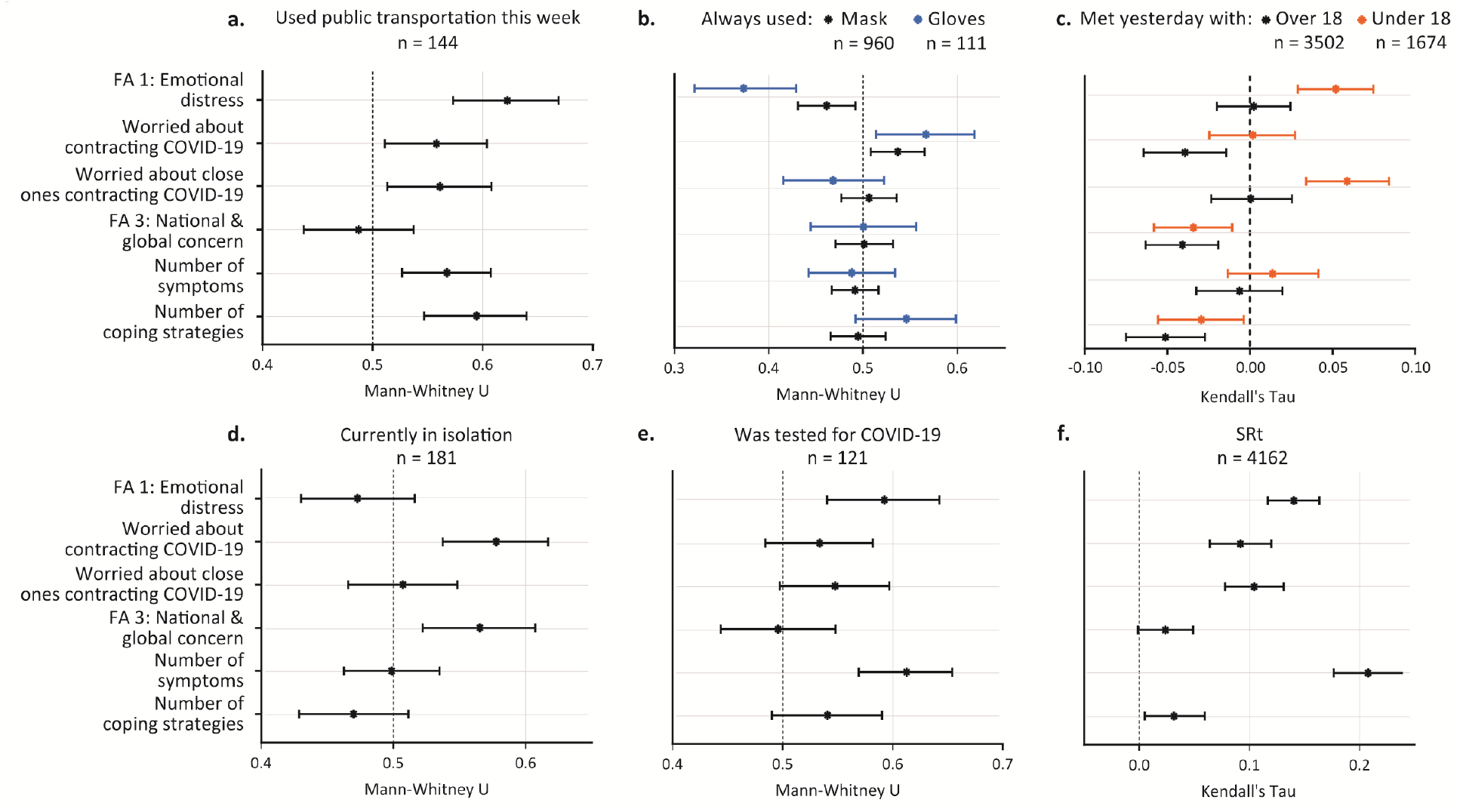
COVID-19 symptoms and related behavioural factors associate with reported distress. (a-f) Effect sizes and 95% confidence intervals of Mann-Whitney’s U (a-b, d-e) and Kendall’s Tau (c, f) for the association between COVID-19-related behaviours and symptoms and respondents’ stress-related responses. SRt: COVID-19 symptoms score.

Second, we found that respondents who reported always wearing gloves, also reported significantly lower levels of general emotional distress (Fig. 8b; n_gloves_always_=111; n_gloves_not_always_=1,397; U=0.37; p=6.3e-5), but a nearly significantly higher level of concern specifically about contracting COVID-19 (Fig. 8b; U=0.57; p=0.077). We found similar trends for mask-wearing, but these effects were slightly smaller (general emotional distress scale: n_mask always_=960; n_mask_not_always_=549; U=0.46; p=0.066; contracting COVID-19: U=0.53; p=0.07). Of note, wearing a mask in Israel has been mandatory since April 4th, while wearing gloves has never been a mandatory requirement, which may explain the difference in both the effect sizes and the sizes of their confidence intervals.

Third, since social relationships are associated with better physical and mental health^29,30^ and better coping with stressful situations^31^, but may also increase the risk of contracting COVID-19, as previously mentioned, we examined whether the number of people respondents had met is associated with their reported distress. Notably, these encounters likely included various types (e.g., meeting family and friends, colleagues or strangers). We found that respondents who reported meeting face-to-face with more people under the age of 18 during the past day reported significantly higher levels of general emotional distress (Fig. 8c; n=4,087; Tau=0.05; p=6.6e-5) and worrying about people close to them contracting COVID-19 (Fig. 8c; Tau=0.06; p=4e-5), but reduced national-global concern (Fig. 8c; Tau=-0.03; p=0.022). In contrast, respondents who reported meeting with more people over the age of 18 did not report higher levels of general emotional distress (Fig. 8c; Tau=0.002; p>0.9), or of worrying about people close to them contracting COVID-19 (Fig. 8c; Tau=5.7e-4; p>0.9). However, they did report significantly lower levels of both worrying about themselves contracting the virus (Fig. 8c; Tau=-0.04; p=0.01) and of national-global concern (Fig. 8c; Tau=-0.04; p=1.5e-3), as well as using fewer stress-coping strategies (Fig. 8c; Tau=-0.05; p=1.8e-4).

We further examined the reports of respondents who are potentially at higher risk for contracting COVID-19, namely, being in quarantine, tested for COVID-19, or experiencing COVID-19 related symptoms. Respondents who reported being in quarantine (due to contact with patients, returning from abroad, having symptoms, or voluntarily isolating oneself) also report significantly higher levels of worrying about contracting COVID-19 (Fig. 8d; n_quarantine_=181; n_not_quarantine_=3,964; U=0.58; p=0.001) and of the national-global concern scale (Fig. 8d; U=0.57; p=0.018). Furthermore, respondents who reported being tested for COVID-19 reported significantly higher levels of general emotional distress (Fig. 8e; n_tested_=120; n_not_tested_=3,967; U=0.59; p=0.003) and experiencing more stress-related symptoms (Fig. 8e; U=0.61; p=3.1e-6).

Finally, respondents who reported experiencing common symptoms of COVID-19 (see *Methods* for the full list of symptoms and the score used here) also reported significantly higher levels of general emotional distress (Fig. 8f; n=4,087; Tau=0.14; p<1e-10), worrying about themselves and their close ones contracting COVID-19 (Fig. 8f; themselves: Tau=0.09; p=3.3e-10; close ones: Tau=0.10; p<1e-10), and more stress-related symptoms (Tau=0.21; p<1e-10). Notably, difficulty breathing may result from both stress and COVID-19 sickness, and therefore appears in both questionnaires, which likely contributes to this strong association between both types of symptoms.

## Discussion

This study explored the behavioural, emotional and mental health impact of the COVID-19 pandemic during six weeks, encompassing the end of the first outbreak and the beginning of the second one in Israel. We used clinically validated instruments (BSI-18, PSS, COPE) to assess symptoms and coping strategies, and questions specifically designed to assess COVID-19-related concerns. As expected, in reaction to stressful events, people reported a variety of concerns, mainly related to their close surroundings (their country and relatives). These non-self-centred concerns may reflect an increased sense of belonging to the country/community. Non-self-centred concerns were reported during the pandemic outbreak in the United States^17^ and also during other times of threat in Israel^32^.

Despite the reported concerns, the anxiety- and depression-related responses (based on the BSI-18 anxiety and depression subscales) are similar to the Israeli norm, based on a nationwide representative sample of 510 community respondents between the ages of 35 and 65 years^14^, and are lower compared with Israelis’ scores during war years^33^. This may be due to sampling at a stage at which the pandemic was mostly well controlled, with low number of COVID-19 cases and deaths. Of note, the normal levels of the anxiety and depression subscales of the BSI assessed in a representative Israeli sample were still higher than the normal levels in the USA and UK at the time^14^. Another important point to consider is that our research sample may not accurately represent the general Israeli population: The percentage of respondents with academic education and the average CSS are higher compared to the general population (70% *vs*. 50.2%, and 187.06 *vs*. 132.7 respectively^34^). Given the negative correlation we found between CSS and emotional distress (Fig. 6a-b), this may explain the relatively low levels of emotional distress in this cohort (Supp. Fig. 1).

Even though levels of general emotional distress were similar to the norm, our study describes the inequalities in mental-health burden associated with the COVID-19 pandemic. As shown, higher emotional burden is associated with being female, younger, unemployed, and living in places with low socioeconomic status. Our findings add to the reported gender differences that were assessed during the COVID-19 outbreak phase in China and in the United States^4,17^. It is interesting to compare our findings to those of the UK COVID-19 social study^35,36^: The number of COVID-19 cases during the outbreak was much higher in the UK compared to Israel; therefore, it is not surprising that contracting COVID-19 remains the most prevalent concern in the UK, and was a lesser concern in Israel. However, despite these differences, in both populations being younger and having lower socioeconomic status correlated with increased emotional distress. The level of community resources may influence individuals’ ability to cope with life challenges, and low socioeconomic places of living, characterized by fewer resources, were found to be more vulnerable in Israel^37,38^ in agreement with the conservation of resources theory^39^.

Employment instability can have devastating effects on the psychological, economic, and social well-being of individuals and communities^40,41^. Despite the importance of this topic during crises^42^, many open questions remain, especially in the context of the COVID-19 pandemic. In our research sample, the people who lost their job due to the pandemic were similar in their levels of emotional distress to those who were unemployed since before the pandemic (Fig. 5a-d). The utilization of more coping strategies by the newly unemployed (Fig. 5e) may also reflect higher levels of distress in this group, which are not shared by those who were unemployed before the pandemic.

Throughout our analysis, we observe a high accordance between the levels of emotional distress, the number of stress-related symptoms, and the number of stress-coping strategies (Figures 3a, 4a, 6a, 7a, 8b,f). This may suggest that employing multiple stress-coping strategies is a sign of inefficient coping, and/or that using multiple strategies is characteristic of highly emotionally disturbed individuals. It would be interesting to investigate whether assessing the coping strategies people use could be used to predict their level of distress.

In summary, although we cannot say what is considered a “normal” response to this new reality, in our research sample the prevalent emotional response to the pandemic was low compared to previous challenging times in Israel. Still, our findings highlight the importance of biological and environmental differences for understanding individuals’ ability to cope with the challenges posed by the pandemic. Such considerations should inform planning and policy for the following waves of the pandemic. In light of the dramatic increase in COVID-19 cases and the unprecedented social-economic crisis that Israel and the rest of the world are experiencing, it is of great importance to continue to investigate the long-term mental-health effects of the pandemic and its consequences.

## Methods

### Online survey

This study used a two-stage online questionnaire, each of which may be considered as a separate one. The first questionnaire was previously described in detail^43^. Briefly, in the first stage, respondents reported on COVID-19-related physiological symptoms (described in the COVID-19 symptoms section) and behaviours (Fig. 8), as well as background demographic (e.g., gender, age and city/town of residence) and medical information (Fig. 7). The latest version of this questionnaire can be access using the following URL: coronaisrael.org

In the second stage, respondents reported on the effects of COVID-19 on their psychological and emotional well-being. These questions were partly based on the anxiety and depression subscales of the brief symptom inventory 18 (BSI-18) and the perceived stress scale (PSS; see full survey description in the methods section and Supp. Fig. 2). Additional questions were designed to assess reasons for concern specifically related to the COVID-19 pandemic, specific stress-related physiological symptoms experienced and stress-coping strategies taken from the brief-COPE questionnaire^15^. Responses were collected in six common languages in Israel (Hebrew, Arabic, English, Russian, French and Spanish), but since very few responses used languages other than Hebrew, only Hebrew responses were analyzed here. The latest version of this questionnaire can be access using the following URL: [forms.gle/4yoXxA3UBuC8R2L68](https://docs.google.com/forms/d/e/1FAIpQLSd9HQDsN1WaikNr0QVrdnG7dKWuevbxR2L1v8Uh62mTHM6c4A/viewform)

## The questionnaire

### The effects of the Coronavirus on the Israeli public

By completing the following questionnaire in full and clicking “submit”, you agree to forward your answers to the questionnaire to Prof. Alon Chen’s research group at the Weizmann Institute of Science.

The Weizmann Institute of Science is a scientific research institution and therefore the study is not intended for clinical purposes or for clinical diagnosis. Filling out this questionnaire will allow crosschecking your answers to the two questionnaires in order to promote the goals of both studies, all according to the privacy policy:

www.weizmann.ac.il/pages/privacy-policy

And the participant form:

www.alonchenlab.com/wp-content/uploads/2020/04/CoronaSurvey_Info.pdf

Please note that the integrated information may identify you to some extent.

Answering this questionnaire is not a substitute for counselling, diagnosis or professional treatment. If you need any of those, we suggest contacting:

Sahar - Network Assistance & Listening - sahar.org.il

Hotline for emergency mental help 1201

Hotline for trauma suffers 1800-363-363

The following sentences refer to your general feelings in the past day. In every statement, please indicate which option most accurately describes your feelings.

I felt irritated:

Not at all Slightly Moderately Largely Very largely

I felt hopeless:

Not at all Slightly Moderately Largely Very largely

I felt tired and restless:

Not at all Slightly Moderately Largely Very largely

I felt scared or anxious:

Not at all Slightly Moderately Largely Very largely

I felt so depressed that nothing could cheer me up:

Not at all Slightly Moderately Largely Very largely

I felt that every task takes so much energy:

Not at all Slightly Moderately Largely Very largely

I felt worthless:

Not at all Slightly Moderately Largely Very largely

I felt lonely:

Not at all Slightly Moderately Largely Very largely

In addition, in the past day, have you experienced the following (check all that apply)?

Increased heart rate

Increased sweating

Trouble sleeping

Loss of appetite

Increased appetite

Difficulty breathing

None of these are true

In every statement, please indicate which option most accurately describes your feelings.

In the past day, to what extent did you feel unable to deal with important things in your life?

Not at all Slightly Moderately Largely Very largely

In the past day, to what extent did you feel confident in dealing with your personal problems?

Not at all Slightly Moderately Largely Very largely

In the past day, to what extent did you feel things are under your control?

Not at all Slightly Moderately Largely Very largely

To what extent, did you feel that you couldn’t cope with challenges facing you?

Not at all Slightly Moderately Largely Very largely

In every statement, please indicate which option most accurately describes your feelings.

I am worried about contracting the coronavirus.

Not at all Slightly Moderately Largely Very largely

I am worried about people close to me contracting the coronavirus.

Not at all Slightly Moderately Largely Very largely

I am worried about my financial situation.

Not at all Slightly Moderately Largely Very largely

I am worried about the situation in Israel.

Not at all Slightly Moderately Largely Very largely

I am worried about the situation around the world.

Not at all Slightly Moderately Largely Very largely

How do the following statements reflect your coping with the situation in the past day (check all that apply):

I tried to accept the situation and learn to live with it.

I used alcohol or cigarettes in order to relax

I used a prescription drug to relax

I drew strength from belief in G-d.

I contacted a family member or friend for support.

I contacted a professional for support.

I searched for information regarding the situation.

I exercised / did yoga/meditation

I drew strength from my pets.

Other

Did you work before the corona pandemic?

I did not work before the corona pandemic.

I worked as a salaried employee.

I worked as a freelancer.

I am retired.

Other

If you worked before the pandemic, what is the status of your employment now?

I am still working

I am on unpaid leave.

I am on paid leave.

I was fired/retired following Covid-19

What is your level of education?

Less than 12 years of study.

High school diploma (Bagrut).

Technical certificate.

Bachelor’s degree and above.

My Gender:

Female

Male

Other

My Age (in years):______________

I live in: ______________

Anything else you would like to add? ______________

### Research sample

Our online questionnaire was made publicly available to anyone with the url, which was posted and distributed using social media starting on April 28th. For this study we used data collected until June 9^th^ 2020. During this period, we collected 12,125 responses from 4,933 respondents. The instructions clearly stated that the questionnaire was intended for adult (18 years old or above) respondents only, and the 74 respondents who indicated they were less than 18 years old were discarded from all of the analyses.

### Note regarding non-binary genders

Respondents were asked to select for their gender either ‘Male’, ‘Female’ or ‘Other’, but since only eight respondents chose ‘Other’, their responses were disregarded in the gender analyses.

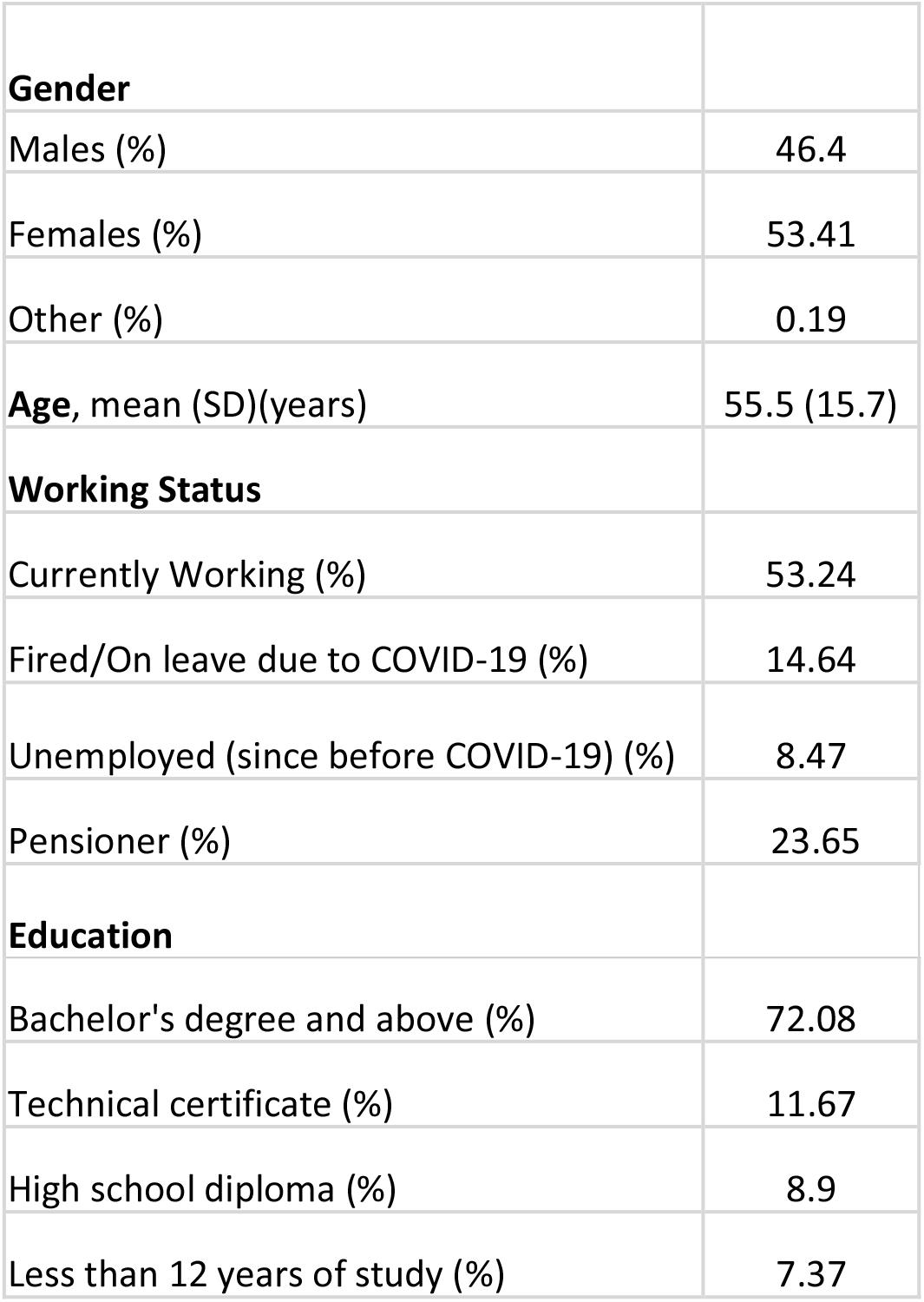

|  |  |
| --- | --- |
| **Gender** |  |
| Males (%) | 46.4 |
| Females (%) | 53.41 |
| Other (%) | 0.19 |
| **Age, mean (SD)(years)** | 55.5 (15.7) |
| **Working Status** |  |
| Currently Working (%) | 53.24 |
| Fired/On leave due to COVID-19 (%) | 14.64 |
| Unemployed (since before COVID-19) (%) | 8.47 |
| Pensioner (%) | 23.65 |
| **Education** |  |
| Bachelor's degree and above (%) | 72.08 |
| Technical certificate (%) | 11.67 |
| High school diploma (%) | 8.9 |
| Less than 12 years of study (%) | 7.37 |

### Categorizing questions using factor analyses

Correlations between individual question responses were quantified using Spearman’s rank correlation coefficient. The resulting correlation matrix was used to compute the factor loadings matrix of a common factor analysis model using the ‘factoran’ Matlab function with the ‘promax’ rotation method. A model of three common factors was chosen using both visual inspection of the factor loading matrices corresponding to a range of models with different numbers of common factors, along with the block diagram of the correlation matrix, and using the ‘nScree’ function of the ‘nFactors’ package in R^44^.

### Statistical analyses

Statistical analyses were performed using Matlab (v.2019b, MathWorks) and R (v-4.0.2). All statistical hypothesis tests are two-tailed and non-parametric, not assuming normality of the marginal or joint distributions of the variables. Bonferroni’s correction for multiple comparisons was used whenever multiple comparisons were made. Unless explicitly stated otherwise, statistical analyses were done based on each respondent’s first response, to ensure the basic assumption of independent samples. The term “statistically significant” was used when p<0.05, following the common convention, but p-values are always shown rounded by at most 5e-4.

For the Mann-Whitney statistic, we denote U=Pr[X<Y]+0.5 Pr[X=Y], where X and Y are randomly chosen observations from the two distributions. Thus, U=1 and U=0 represent complete separation of the distributions, while U=0.5 represents complete overlap. The U statistic and its corresponding p-values and confidence intervals were calculated using the ‘wmwTest’ function of the ‘asht’ package in R^45^. Odds ratios and their corresponding p-values and confidence intervals were calculated using the ‘oddsratio’ function of the ‘epitools’ package in R^46^ with Fisher’s exact conditional maximum likelihood estimation option. The Kendall rank correlation coefficient (commonly known as Kendall’s Tau) and its corresponding p-values were calculated using the Matlab ‘corr’ function and its corresponding confidence intervals were calculated using bootstrapping with the Matlab ‘bootci’ function and the ‘bias corrected and accelerated percentile’ method^47,48^.

### Visualizing the association between two ordinal variables

Since both our independent and dependent ordinal variables (e.g., stress-related responses, age, city socioeconomic status) had numerous repeated values, heatmaps were used instead of scatter plots to visualize their relationships. The data were divided into five equally numerous bins whenver possible (i.e., whenever distinct quintiles existed). Otherwise, wherever no distinct quintiles existed (e.g., for the number of reported stress-related symptoms in Fig. 3e), the raw values were used. We chose to use five bins whenever possible to match the number of possible responses in most of our questions. Under the null hypothesis – that the variables are statistically independent – all heatmap cells are expected to have roughly the same frequency (f = n/d, where n = number of responses and d = number of heatmap cells), which makes it very straightforward to examine visually. Importantly, the correlation coefficients and the corresponding p-values and confidence intervals were quantified using the raw data, i.e., without any binning (see ‘Methods - Statistical analyses’).

### COVID-19 symptoms score (SRt)

The COVID-19 symptoms score used here was previously described in detail^49^. Briefly, this score aims to reflect the importance of each symptom with respect to its prevalence in confirmed COVID-19 patients, as previously reported^50^. The symptoms included in the score calculation were: fever (79% of confirmed COVID-19 patients), shortness of breath (3.5%), cough (58%), fatigue (29.3%), muscle pain (3.8%), sore throat (3.2%), headache (6%) and diarrhea (5.7%).

## Data Availability

To protect the privacy of the study participants and their sensitive information (e.g., medical information, street address, and other personal information), the whole dataset and the code that processes it will not be made publicly available as-is, but specific data and code, which may be needed for reproducing results, will be made available by the corresponding author upon reasonable request.

## Acknowledgements

The authors wish to thank Dr. Jessica Keverne for writing and editing support and advice. A.C. is the incumbent of the Vera and John Schwartz Professorial Chair in Neurobiology at the Weizmann Institute of Science; the Head of the Max Planck Society–Weizmann Institute of Science Laboratory for Experimental Neuropsychiatry and Behavioral Neurogenetics gratefully funded by the Max Planck Foundation; and the Head of Ruhman Family Laboratory for Research in the Neurobiology of Stress at the Weizmann Institute of Science. This work is supported by The Weizmann Institute Coronavirus Response Fund (A.C. and E.S); research support from Bruno and Simone Licht (A.C.) and Roberto and Renata Ruhman (A.C.). Y.K. is the incumbent of the Sarah and Rolando Uziel Research Associate Chair.

