## Supplementary figures 1-5 for "Stress-related emotional and behavioural impact following the first COVID-19 outbreak peak"

### SUPPLEMENTARY MATERIAL

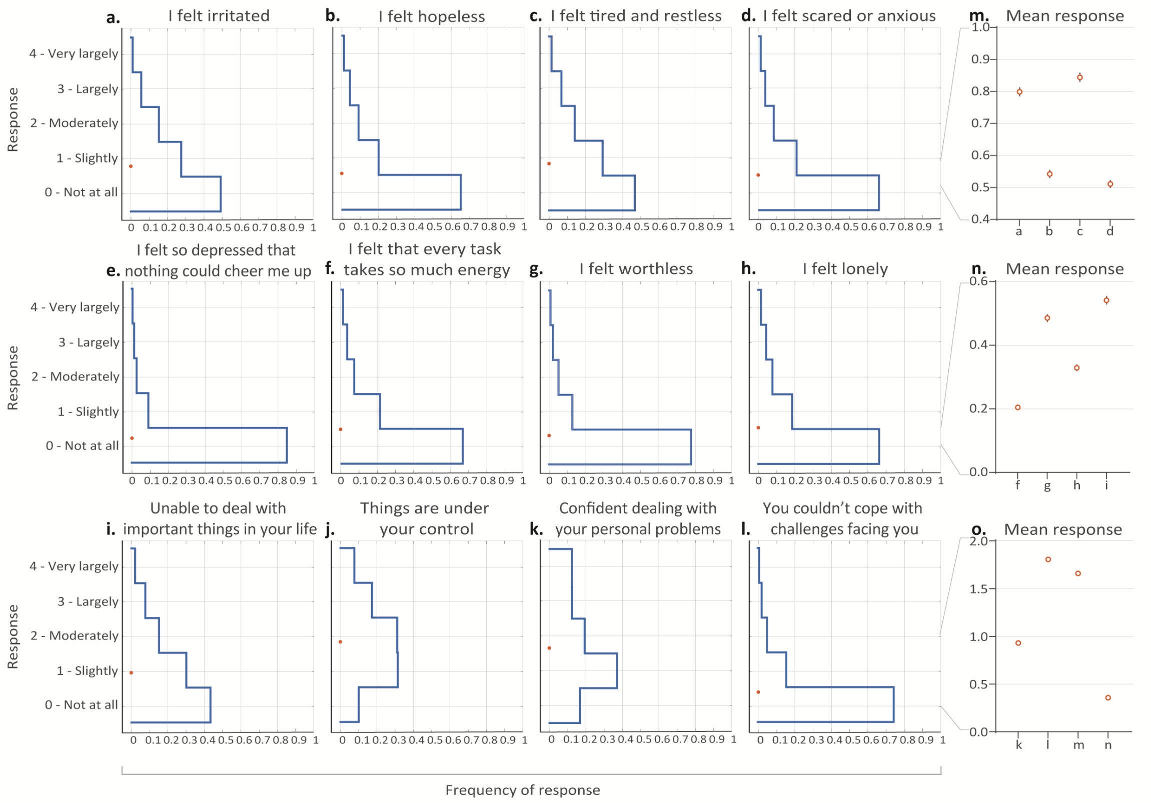


**Supplementary Figure 1. Stress-related response distributions.** (a-e, f-i, k-n) Distributions of responses to emotional distress questions among all respondents. Red circles represent response means. (e, j, o) Zoomed-in view of the response means shown in panels a-e, f-i, k-n.

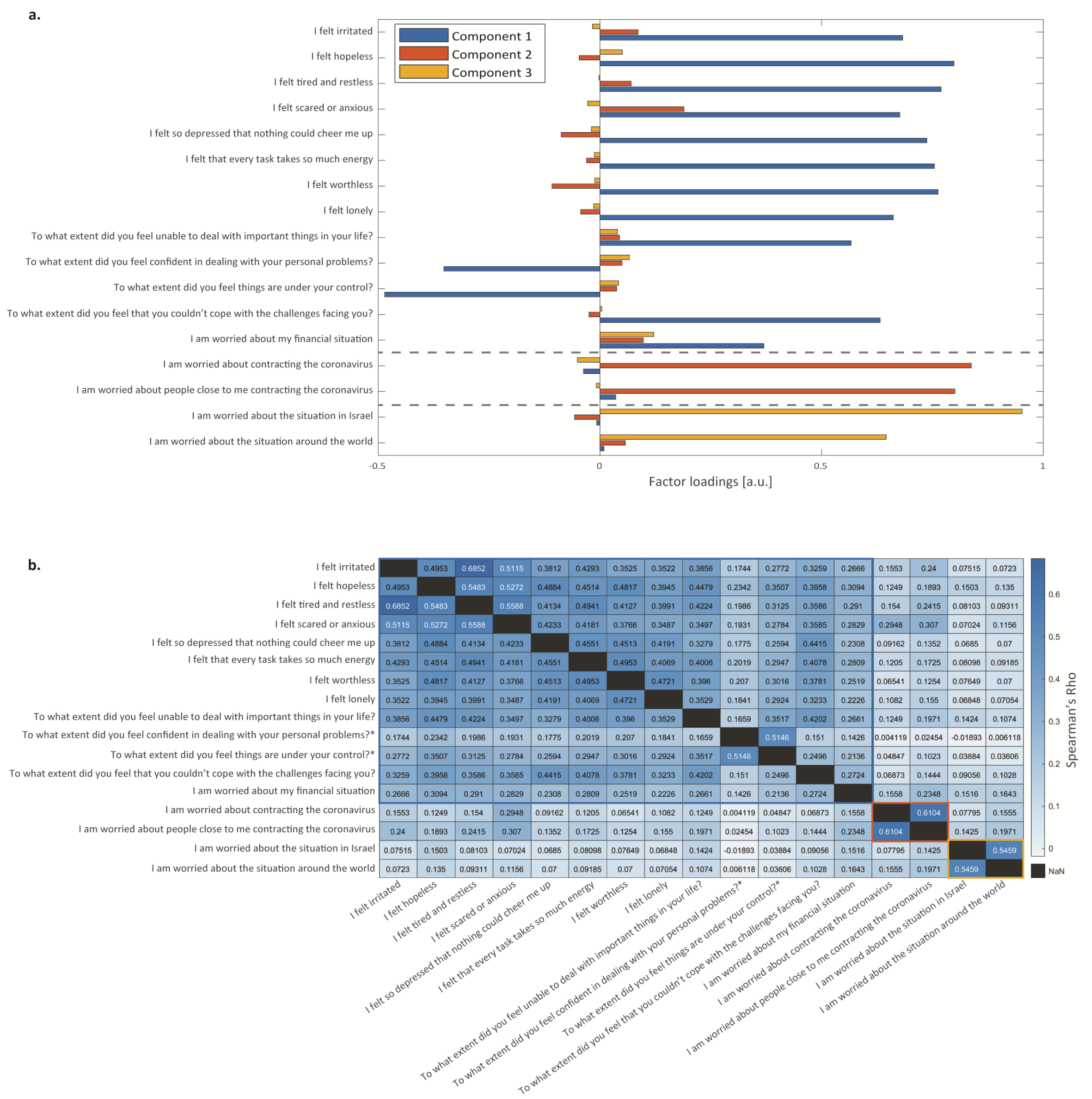


**Supplementary Figure 2. Categorizing stress-related questions using factor analysis.** (a) Factor loadings of individual stress-related responses onto the three main factors inferred using factor analysis. (b) Correlation matrix showing the correlation between each pair of questions. The block diagonal structure demonstrates the division into question subsets. To visualize the correlation more clearly, the ordering of the responses of two questions (marked with an asterisk in the figure) was reversed (i.e., 'Not at all' = '5' and 'Very largely' = 1) during this analysis, to match the "valence" of the other questions: "To what extent did you feel confident in dealing with your personal problems" and "To what extent did you feel things are under your control".

| Did not report | | Did report | | | |
|---|---|---|---|---|---|
| F | M | F | M | P-val | |
| 1776 | 1711 | 749 | 501 | 7.26e-07 | Trouble sleeping |
| 2279 | 2069 | 246 | 143 | 0.00063 | Increased appetite |
| 2376 | 2133 | 149 | 79 | 0.00263 | Increased heartrate |
| 2410 | 2131 | 115 | 81 | 0.86604 | Loss of appetite |
| 2434 | 2155 | 91 | 57 | 0.49598 | Difficulty breathing |
| 2455 | 2174 | 70 | 38 | 0.24794 | Increased sweating |
| 907 | 811 | 1618 | 1401 | 0.99999 | I tried to accept the situation and learn to live with it |
| 1252 | 1193 | 1273 | 1019 | 0.04344 | I exercised, did yoga or meditated |
| 1472 | 1346 | 1053 | 866 | 0.69043 | I searched for information regarding the situation |
| 2079 | 2054 | 446 | 158 | < 1e-10 | I contacted a family member or friend for support |
| 2150 | 2008 | 375 | 204 | 4.48e-08 | I drew strength from my pets |
| 2275 | 2005 | 250 | 207 | 0.99999 | I drew strength from belief in God |
| 2367 | 2094 | 158 | 118 | 0.95894 | I used alcohol or cigarettes in order to relax |
| 2391 | 2121 | 134 | 91 | 0.57504 | I used a prescription drug to relax |
| 2443 | 2172 | 82 | 40 | 0.02569 | I contacted a professional for support |

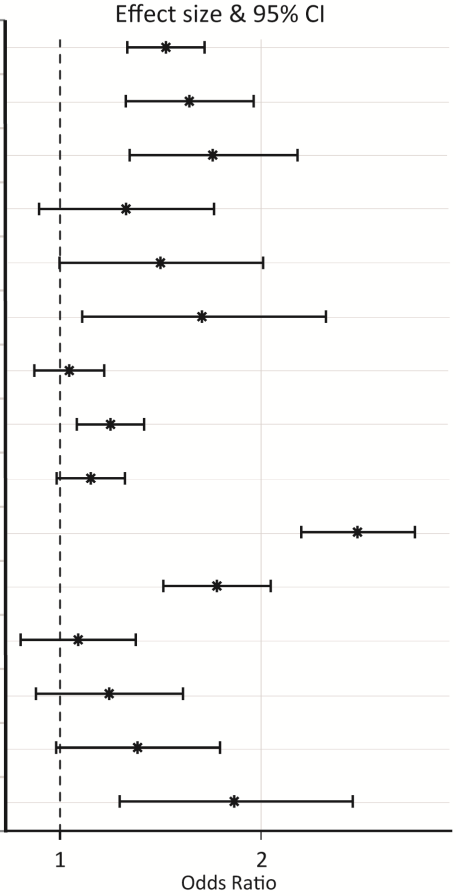


**Supplementary Figure 3. Association between gender and stress-related symptoms and coping strategies.** Left: Table includes the number of positive and negative answers for women and men, as well as the p-values for the association between gender and each stress-related symptom and coping strategy. Right: Odds Ratio and 95% confidence intervals for female vs. male responses on each variable.

| Did not report | Did report | P-val | |
|---|---|---|---|
| 3479 | 1249 | 5.71e-08 | Trouble sleeping |
| 4338 | 390 | 2.52e-06 | Increased appetite |
| 4500 | 228 | 1.10e-06 | Increased heartrate |
| 4532 | 196 | < 1e-10 | Loss of appetite |
| 4580 | 148 | 0.08536 | Difficulty breathing |
| 4620 | 108 | 2.68e-05 | Increased sweating |
| 1718 | 3010 | 0.50062 | I tried to accept the situation and learn to live with it |
| 2442 | 2286 | < 1e-10 | I exercised, did yoga or meditated |
| 2812 | 1916 | 0.13584 | I searched for information regarding the situation |
| 4125 | 603 | < 1e-10 | I contacted a family member or friend for support |
| 4152 | 576 | 5.64e-08 | I drew strength from my pets |
| 4272 | 456 | < 1e-10 | I drew strength from belief in God |
| 4452 | 276 | < 1e-10 | I used alcohol or cigarettes in order to relax |
| 4503 | 225 | > 1-1e-10 | I used a prescription drug to relax |
| 4606 | 122 | < 1e-10 | I contacted a professional for support |

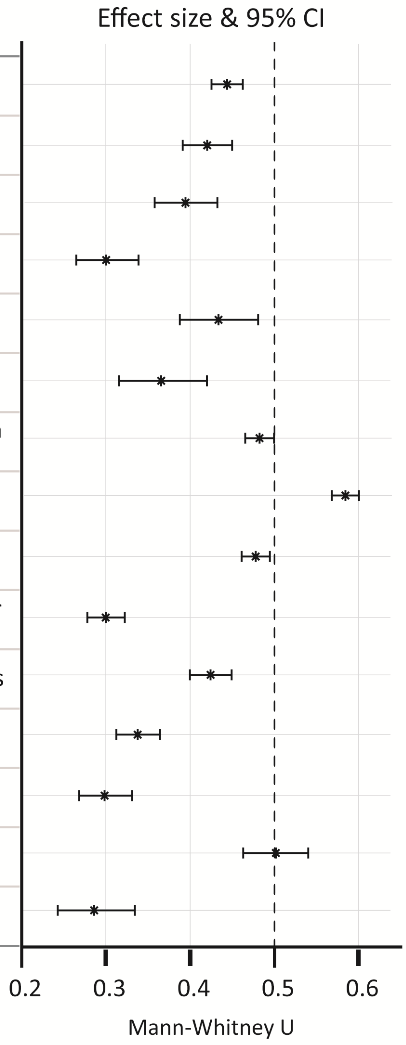


**Supplementary Figure 4. Association between age and stress-related symptoms and coping strategies.** Left: Table includes the number of positive and negative answers, as well as the p-values for the association between age and each stress-related symptom and coping strategy. Right: Mann-Whitney U and 95% confidence intervals for each variable.

| Did not report | Did report | P-val | |
|---|---|---|---|
| 2982 | 1106 | 0.92715 | Trouble sleeping |
| 3756 | 332 | > 1-1e-10 | Increased appetite |
| 3880 | 208 | 0.99995 | Increased heartrate |
| 3901 | 187 | 0.19977 | Loss of appetite |
| 3954 | 134 | 0.037233 | Difficulty breathing |
| 3990 | 98 | > 1-1e-10 | Increased sweating |
| 1450 | 2638 | 0.9998 | I tried to accept the situation and learn to live with it |
| 2130 | 1958 | 0.00125 | I exercised, did yoga or meditated |
| 2406 | 1682 | 0.99992 | I searched for information regarding the situation |
| 3553 | 535 | 0.00509 | I contacted a family member or friend for support |
| 3600 | 488 | 0.06136 | I drew strength from my pets |
| 3686 | 402 | < 1e-10 | I drew strength from belief in God |
| 3830 | 258 | 0.97593 | I used alcohol or cigarettes in order to relax |
| 3885 | 203 | 0.38714 | I used a prescription drug to relax |
| 3977 | 111 | > 1-1e-10 | I contacted a professional for support |

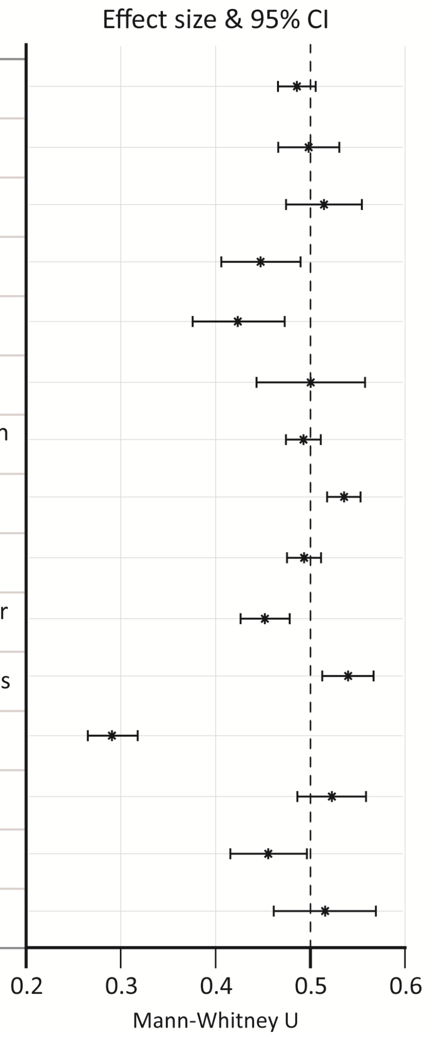


**Supplementary Figure 5. Association between city socioeconomic status and stress-related symptoms and coping strategies.** Left: Table includes the number of positive and negative answers, as well as the p-values for the association between socioeconomic status and each stress-related symptom and coping strategy. Right: Mann-Whitney U statistic and 95% confidence intervals for each variable.
